# Integrated chronic disease care delivery at a primary care level in sub-Saharan Africa: A systematic review, ‘best fit’ framework synthesis, and new conceptual model

**DOI:** 10.1101/2021.11.30.21267057

**Authors:** Aileen Jordan, Simon Harrison

## Abstract

**Background:** Existing chronic care conceptual models were not designed for sub-Saharan Africa, where there is a growing burden of chronic disease. This review provides a qualitative synthesis and new conceptual model for primary care approaches to the integration of chronic communicable and non-communicable disease care in a sub-Saharan African context.

**Methods:** A ‘best fit’ framework synthesis comprising two systematic reviews, with information retrieved from PubMed, Embase, CINAHL plus, Global Health, and Global Index Medicus databases between 1^st^ – 30^th^ April 2020. Articles on chronic care conceptual models were included if they were developed for application in a primary care context and described a framework for long-term management of chronic disease care, and themes extracted to construct an *a priori* framework. A second systematic review included articles focussing on integrated HIV and diabetes care at a primary care level in sub-Saharan Africa, with thematic analysis carried out against the *a priori* framework. A new conceptual model was constructed from *a priori* themes and new themes. Risk of bias of included studies was assessed using CASP and MMAT.

**Results:** Two conceptual models of chronic disease care, comprising 6 themes, were used to build the *a priori* framework. The systematic review of primary research identified 12 articles, with all 6 of the *a priori* framework themes, and 5 new themes identified. A new patient-centred conceptual model for integrated HIV and diabetes care was constructed, specific to a sub-Saharan African context.

**Discussion:** Improving patient access to chronic disease care through implementing decentralised, integrated, affordable and efficient primary care services should be prioritised in sub-Saharan Africa. Services must be acceptable to patients, viewing them as partners, addressing their concerns, and seeking to safeguard confidentiality. Limitations of this study include potential publication bias, and impact of policy environment and economic factors in sub-Saharan Africa.

**SUMMARY BOX:** *What is already known?:*

- The health transition taking place in sub-Saharan Africa (SSA) towards chronic communicable and non-communicable diseases such as HIV and diabetes as the main causes of morbidity and mortality means that health systems currently orientated towards acute, episodic care, must be re-orientated towards meeting the long-term needs of patients with chronic diseases.
- Existing chronic care conceptual models were designed for use in high income countries rather than a SSA context.

*What are the new findings?:*

- All 6 of the *a priori* framework themes derived from the Chronic Care Model and the ICCCF were identified within the primary research studies and therefore have relevance to the provision of chronic care in a primary care context in SSA.
- An additional 5 new themes were identified from the primary research studies; improving patient access, task-shifting, clinical mentoring, stigma and confidentiality, and patient-provider partnerships.

*What do the new findings imply?:*

- These findings imply that there are additional themes and delivery strategies specific to an SSA context that need to be considered in the implementation of primary care level integrated chronic disease care provision in SSA.
- The new themes identified from the primary research highlight the importance of health services being accessible and acceptable to patients, of partnering with patients to improve health outcomes, and of patient confidentiality and imply a need to reconceptualise chronic care from a patient-centred viewpoint.

## INTRODUCTION

Sub-Saharan Africa (SSA) is experiencing an epidemic of non-communicable disease^1^, with deaths from non-communicable disease projected to be greater than those resulting from communicable, perinatal, maternal and nutritional causes combined by 2030^2 3^. However, SSA populations also remain at risk from communicable diseases such as HIV, which continues to be a leading cause of morbidity and mortality in SSA^4 5 6 7^. Both chronic non-communicable diseases such as diabetes and chronic communicable diseases such as HIV require sustained engagement with healthcare services for long-term follow-up and treatment^8 9 10 11 12 13^. HIV/AIDS is the top cause of Disability-adjusted life years (DALYs) lost within the adult population in SSA; diabetes is of rising concern and in 2019 was the third most common cause of DALYs lost due to non-communicable disease in over 50 year olds in SSA^6^. Therefore, one of the key strategic challenges facing health policy makers in SSA is how fragmented health systems, traditionally orientated towards addressing acute episodic care, can be strengthened to provide continuity of care and address both chronic communicable and non-communicable diseases^8^.

This double burden of disease in SSA from both chronic communicable and non-communicable disease means that there is an urgent need to move beyond vertical interventions for individual diseases, towards an integrated approach to healthcare provision for chronic disease of all causes^8 14^. Integrated primary care has the goal of sustainably improving quality and continuity of care for people with chronic conditions^15^, and building strong primary care-based systems has been recognised by WHO as a key strategic goal in the provision of integrated, people-centred health services^10 16 17^. In high income countries an integrated approach to chronic disease management in primary care has been extensively implemented based upon the chronic care model (CCM) developed by Wagner *et al*.^18 15^. However, as yet, there is no consensus on a model to guide implementation of integrated primary care which is specific to the unique challenges of providing chronic disease care in low- and middle-income countries (LMICs) in SSA^15 19^.

The objective of this study is to describe a model of primary care that addresses the integrated management of both chronic communicable and non-communicable disease in a SSA context. Due to their high burden of disease, diabetes and HIV have been chosen as examples of non-communicable and communicable chronic diseases that exemplify the challenges of delivering high-quality long-term chronic disease care to patients in SSA.

## METHODS

The ‘best fit’ framework synthesis (BFFS) method was selected as it provides a systematic and transparent method to identify and build upon existing frameworks and models of chronic disease care using primary research data to derive a new model relevant to a SSA context^20^. The BFFS method utilises both framework and thematic synthesis approaches to analyse complex issues of feasibility and health system implementation^21^, leading to synthetic products that are reproducible and relevant to policymakers and those designing interventions^22^. The BFFS method has been previously utilised by Lall *et al*. and Kane *et al*. to adapt existing models of chronic disease care for non-communicable diseases in LMIC settings^19 23^.

The 7 steps of the BFFS method were followed for this systematic review20: (1) the review question was formulated; (2) two systematic reviews were conducted (a) to identify existing relevant ‘best fit’ chronic disease care conceptual models and (b) of primary research focusing on integration of HIV and diabetes care at a primary care level in SSA; (3) results of the systematic reviews were then analysed to (a) construct an *a priori* framework using thematic analysis and (b) assess primary research study characteristics and quality; (4) study evidence was coded against the *a priori* framework; (5) any data that could not be placed within the framework was interpreted using inductive, thematic analysis; (6) a new framework was created using both the *a priori* and new themes identified from the primary research; (7) further thematic analysis led to the creation of a new conceptual model.

### Search strategy

#### *A priori* framework

PubMed, Embase, CINAHL plus, Global Health (CABI), and WHO’s Global Index Medicus databases were searched to identify conceptual models or frameworks of chronic disease care for the generation of the *a priori* framework. The search terms for PubMed were Primary health care [MeSH] OR community health services [MeSH] OR delivery of healthcare, integrated [MeSH] OR primary prevention, organisation and administration [MeSH]; AND chronic disease, prevention and control [MeSH] OR chronic disease, therapy [MeSH] OR chronic disease, epidemiology [MeSH] AND models, organisational [MeSH]. Search terms were adapted as appropriate for each database.

Inclusion criteria were articles in English, describing a conceptual model or framework for chronic disease care, developed for application in a primary care context, and describing the long-term management of chronic diseases. We excluded animal models, purely economic models, models describing healthcare workforce roles only, models focusing on a specific aspect of chronic disease care provision such as health promotion, screening, self-care, e-health or telemedicine, or on a population subgroup such as paediatric or elderly care. Models that were an adaption of a pre-existing conceptual model to a specific country or health system context not relevant to SSA were also excluded. No limits were set on date of publication. Database searches were carried out between 1^st^ and 30^th^ April, 2020.

#### Primary research studies

PubMed, Embase, CINAHL plus, Global Health (CABI), and WHO’s Global Index Medicus were searched for primary research studies, along with grey literature via Google Scholar and analysis of key study bibliographies for relevant studies. The validated broad integrated care search filter for PubMed^24 25 26^, was incorporated into the search strategy. The search terms used for PubMed were Primary care [MeSH] OR Community Health Services[MeSH] OR Chronic Care [All fields] OR (Integrated care broad search); AND Diabetes mellitus [MeSH] OR Diabetes [All fields] OR noncommunicable diseases [MeSH] OR noncommunicable diseases [All fields]; AND Human Immunodeficiency Virus [MeSH] OR HIV infections [MeSH] OR HIV [All fields]; AND Africa South of the Sahara [MeSH] OR individually named sub-Saharan African countries.

Articles in English conducted in sub-Saharan African countries, as defined by World Bank Regional Groups, evaluating the implementation of an intervention or programme to integrate HIV and diabetes care located primarily at the primary level of the healthcare service were included^27^. Database searches were carried out between 1^st^ and 30^th^ April 2020. Articles published between September 2010 and 2020 were included, to provide relevant data that would reflect the current SSA primary care context.

### Analysis

Articles retrieved during each search process were imported into bibliographic management software. Duplicate records were removed, followed by an initial screen of titles and abstracts for relevance. Full-text screening was then carried out, recording reasons for exclusion. Both authors screened and confirmed the final selection of articles. Data was extracted from all included articles into custom-made data extraction sheets.

Model elements were extracted from the chronic care models identified and used to build the *a priori* framework themes to which the primary research studies were mapped.

Primary research studies were analysed and coded using the *a priori* framework themes. Following this, the results of primary research studies which could not be mapped to the *a priori* framework were thematically analysed and new themes identified. Data quality of the primary research studies was assessed as part of the systematic review. Studies were appraised against the Critical Appraisal Skills Programme Checklist (CASP)^28^ and the Mixed Methods Appraisal Tool (MMAT)^29^. A study was deemed to be of adequate quality for inclusion if >50% of the CASP and MMAT checklists had been met. All the studies were found to be of adequate quality for inclusion.

## RESULTS

### A priori framework

An initial 422 articles were identified and after removing duplicates, 377 articles were screened by title and abstract with 366 then excluded. The 11 potentially applicable studies describing models or frameworks were then assessed with a further 9 excluded, as outlined in Figure 1.

**Figure 1.**
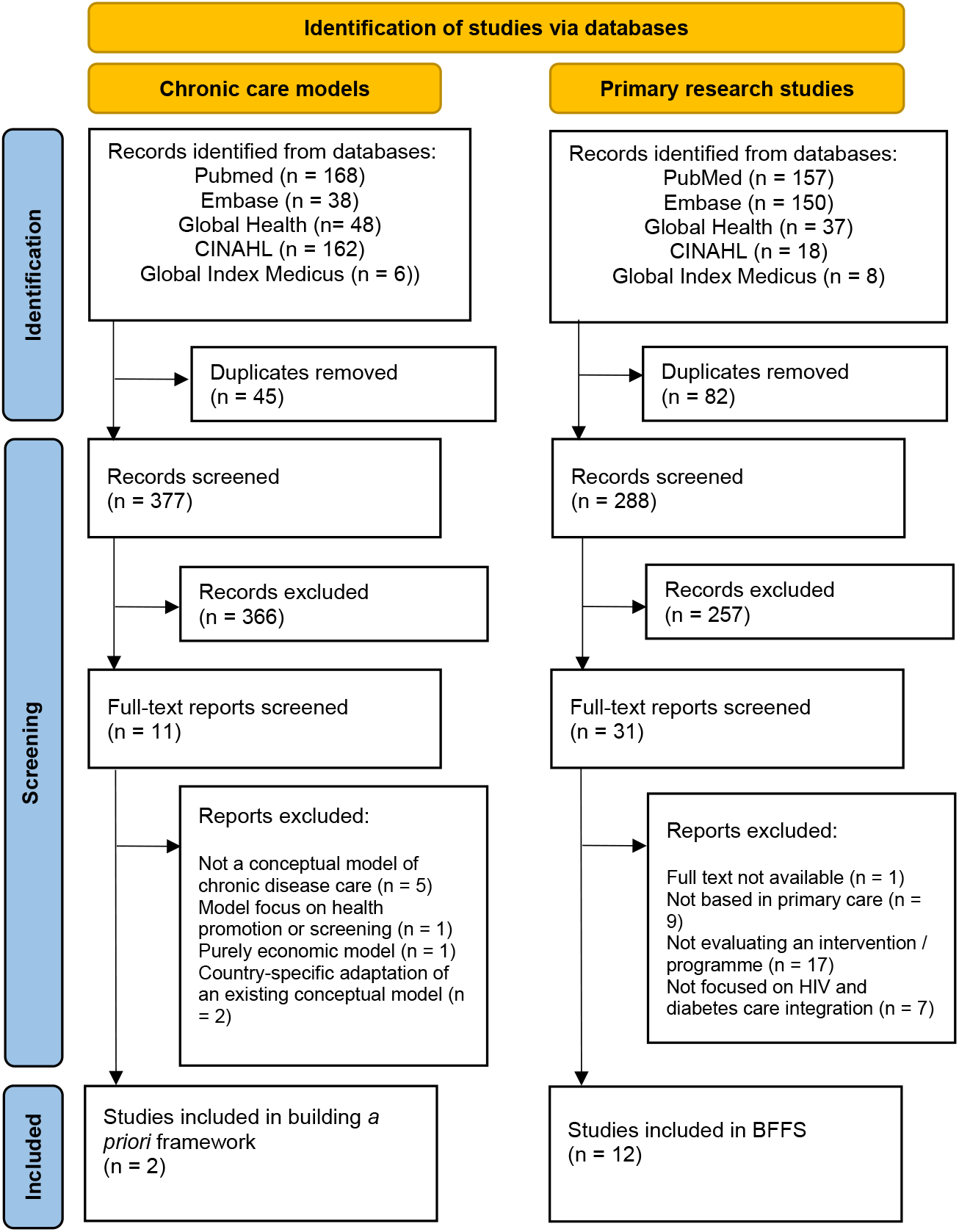
PRISMA flowchart of search results for models used to construct *a priori* framework, and primary research studies used for best fit framework synthesis (BFFS). Adapted from Page *et al*.^72^.

Two models satisfied the inclusion criteria for informing the *a priori* framework; the Chronic Care Model^18^, and the Innovative Care for Chronic Conditions Framework (ICCCF)^30 31^. Model elements and building blocks of these models were extracted to inform development of the *a priori* framework (Table 1). The CCM identifies 6 essential model elements that enable provision of high-quality chronic disease care in a primary care setting^18 32 33^. Each of these model elements can be further described in terms of service delivery components or strategies that enable the implementation of integrated chronic disease care^34^. In 2003 several additional service delivery components were added to reflect pilot study results^35^ and these were included for analysis. The scope of the ICCCF conceptual model is broader than the CCM, describing micro, meso and macro levels of integration^30^. Primary care as a service delivery approach is located at the meso level of integration^36^ therefore the meso level building blocks described in the ICCCF were used to inform the *a priori* framework. Although these meso level building blocks of the ICCCF are based upon the 6 model elements of the CCM and the models overlap significantly in their content^31^, it was felt both the CCM and ICCCF contain unique service delivery strategies which were valuable to inform the *a priori* framework themes.

**Table 1.** A priori framework themes identified from CCM and ICCCF conceptual models^30 32 35^.

| CCM model element | ICCCF building block | Service delivery strategies | *A priori* framework themes |
| --- | --- | --- | --- |
| **Delivery system design** | **Promote continuity and coordination** | Team members have defined roles and responsibilities within team <sup>a</sup><br>Care appropriate to cultural background <sup>a</sup><br>Clinical case management for complex patients <sup>a</sup><br>Promotion of communication within healthcare team to coordinate patient care <sup>b</sup><br>Continuity through planned and proactive follow-up to support evidence-based care <sup>c</sup><br>Coordination of services across levels of healthcare and providers <sup>c</sup> | [1] *Effective team-working to deliver continuity and coordinated proactive care* |
| **Health System** | **Encourage quality care through leadership and incentives** | Promotion of effective strategies for improvement and system change <sup>a</sup><br>Organisational culture and leadership support of improving quality of care <sup>c</sup><br>Incentivisation of quality care <sup>c</sup> | [2] *Organisational leadership, culture and mechanisms to promote quality and safety* |
|  |  | Quality monitoring and improvement activities as a routine among all team members, including processes for review of significant events<sup>c</sup> |  |
| **Decision support** | **Organise and equip healthcare teams** | Promote continuing professional education<sup>a</sup><br>Access to specialist advice<sup>a</sup><br>Equipped health care teams with necessary supplies, medical equipment, laboratory access, and essential medications<sup>b</sup><br>Training to help promote patient self-management and support behavioural change<sup>b</sup><br>Implement ongoing use of evidence-based guidelines and diagnostic and treatment algorithms<sup>c</sup><br>Effective communication to promote information exchange and patient participation in shared decision-making in evidence-based management<sup>c</sup> | [3] *Equipped healthcare teams to deliver evidence-based patient-centred care* |
| **Self-management support** | **Support self-management and prevention** | Proactive support for patient's self-management and prevention efforts over time<sup>b</sup><br>Emphasise central role of patient in managing care<sup>c</sup><br>Utilisation of effective self-management strategies<sup>c</sup><br>Organisation of resources to support patient self-management and prevention<sup>c</sup> | [4] *Empowerment and support of patients for self-management and prevention* |
| **Clinical Information Systems** | **Use information systems** | Monitoring of team performance<sup>a</sup><br>Sharing of information with patients and providers for care coordination<sup>a</sup><br>Collect and organise useful patient data<sup>b</sup><br>Reminder systems for providers and patients<sup>c</sup><br>Identify sub-populations for proactive care<sup>c</sup><br>Use for individual care planning<sup>c</sup> | [5] *Use of data collection systems to facilitate effective care and follow-up* |
| **Community resources and policies** | **Building Blocks for the Community** | Encourage participation of patients in effective community programmes<sup>a</sup><br>Raise awareness of chronic conditions and reduce stigma<sup>b</sup><br>Provide complementary preventive and management services through mobilising informal network of providers, such as community health workers and volunteers<sup>b</sup><br>Mobilise and coordinate local community resources to support screening, prevention, and improved management of chronic conditions<sup>b</sup><br>Form partnerships with community leadership and organisations<sup>c</sup> | [6] *Community partnerships to promote awareness, mobilise resources and support health service provision* |

a - Model element found in CCM only<sup>18</sup>, b - Model element found in ICCCF only<sup>30</sup>, c - Model element found in both CCM and ICCCF<sup>18 30</sup>.

### Primary research studies

Database searching retrieved 370 citations, with 288 unique citations after removing duplicates (Figure 1). 31 full-text articles were screened, of which 12 were found to satisfy the inclusion criteria previously described. There were no additional records identified from searches of bibliographies of key articles or of grey literature using Google Scholar. The 12 studies included in the framework synthesis varied by design, with 7 qualitative studies, 4 quantitative studies, and 1 mixed methods study identified. The 12 studies were conducted in 8 different SSA countries; South Africa (n=3); Kenya (n=3); Malawi (n=2), Uganda (n=1); Botswana (n=1); Zimbabwe (n=1); Eswatini (n=1) and Ethiopia (n=1) (see table 2).

**Table 2.**
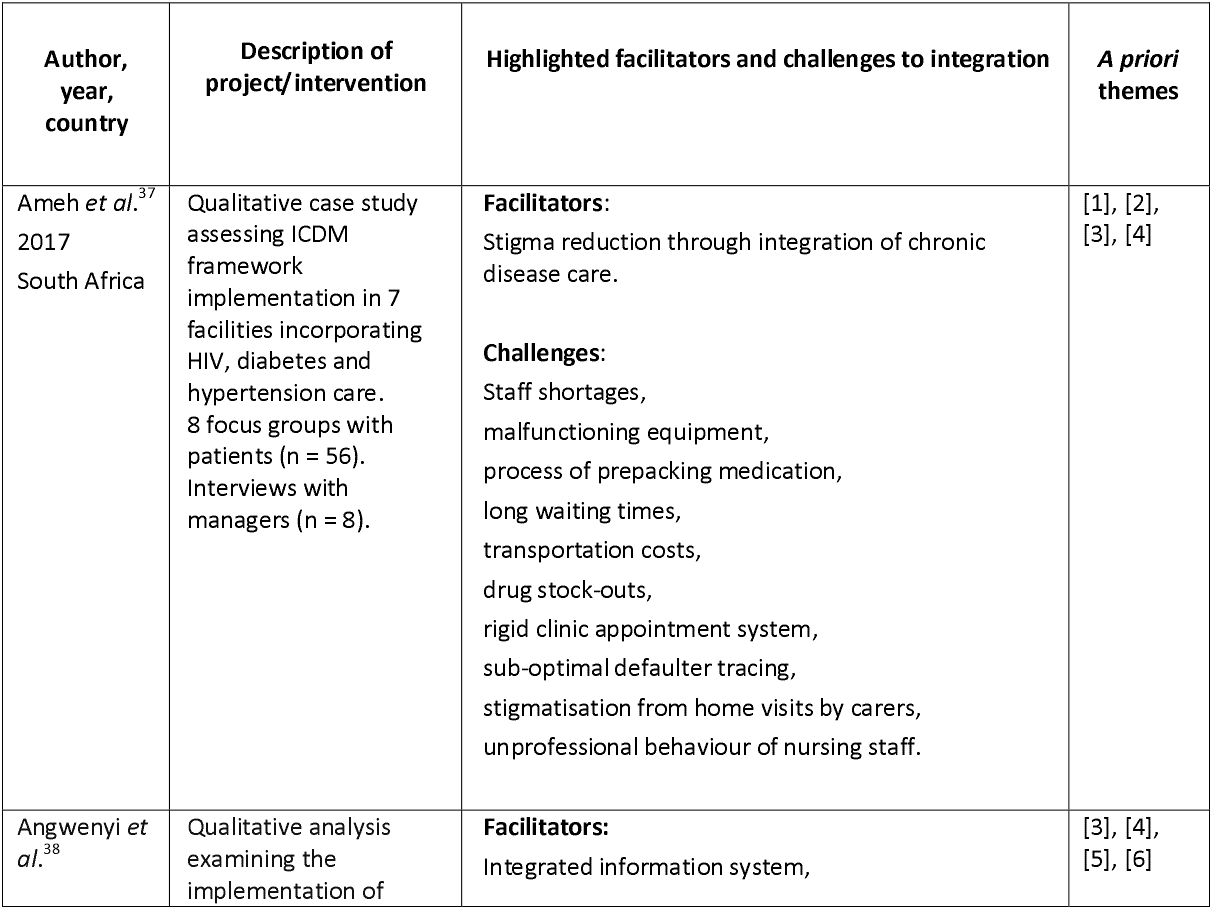
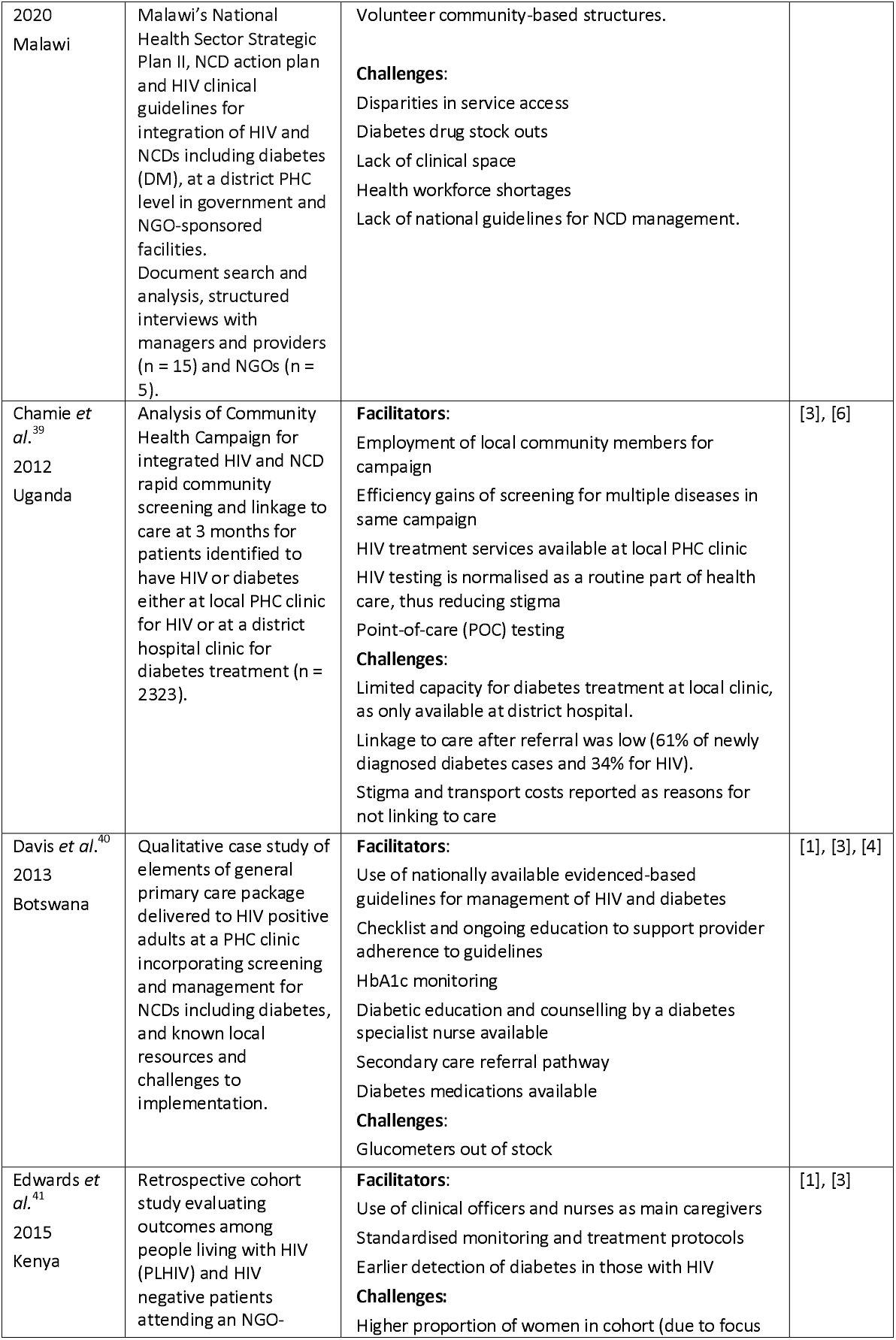
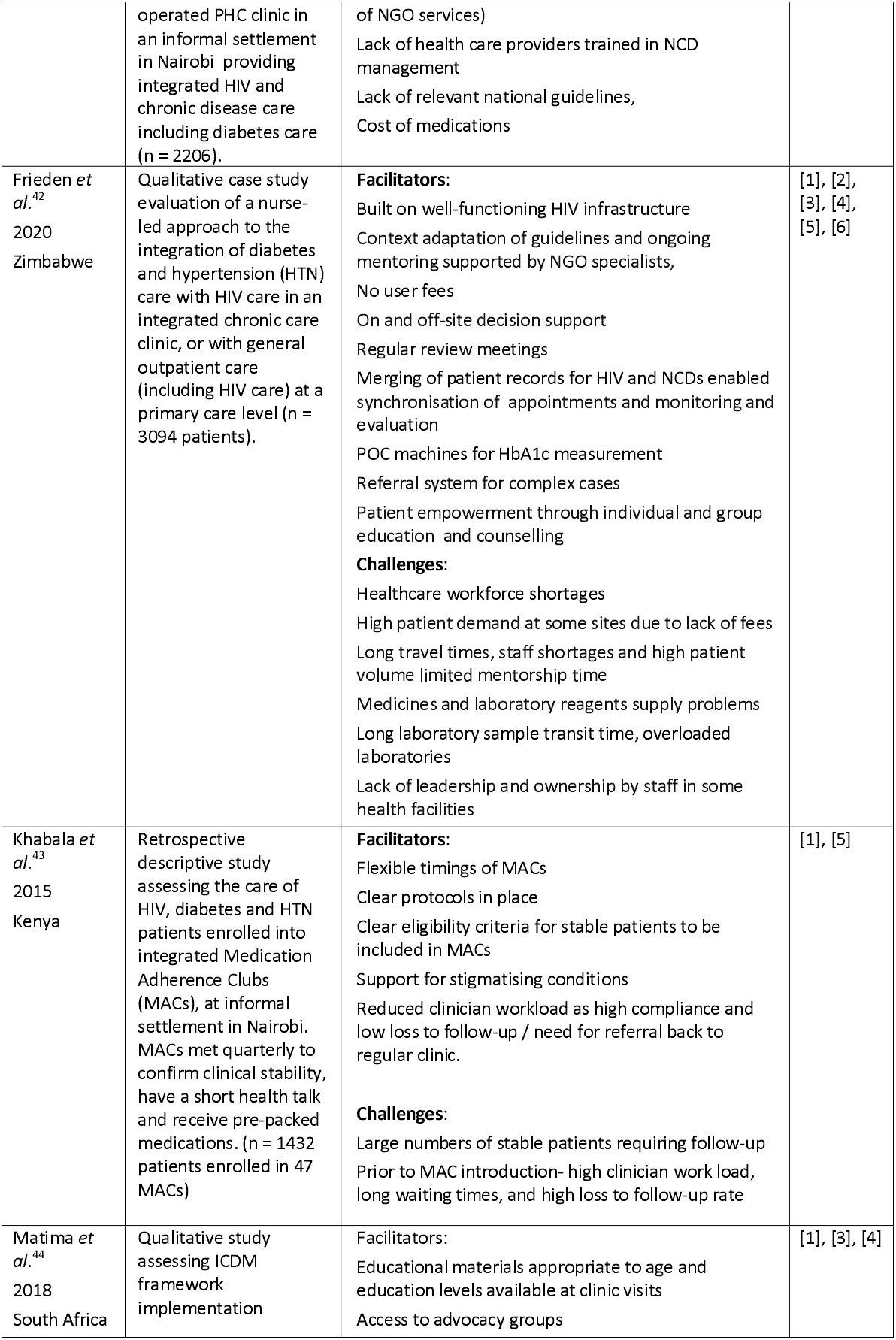
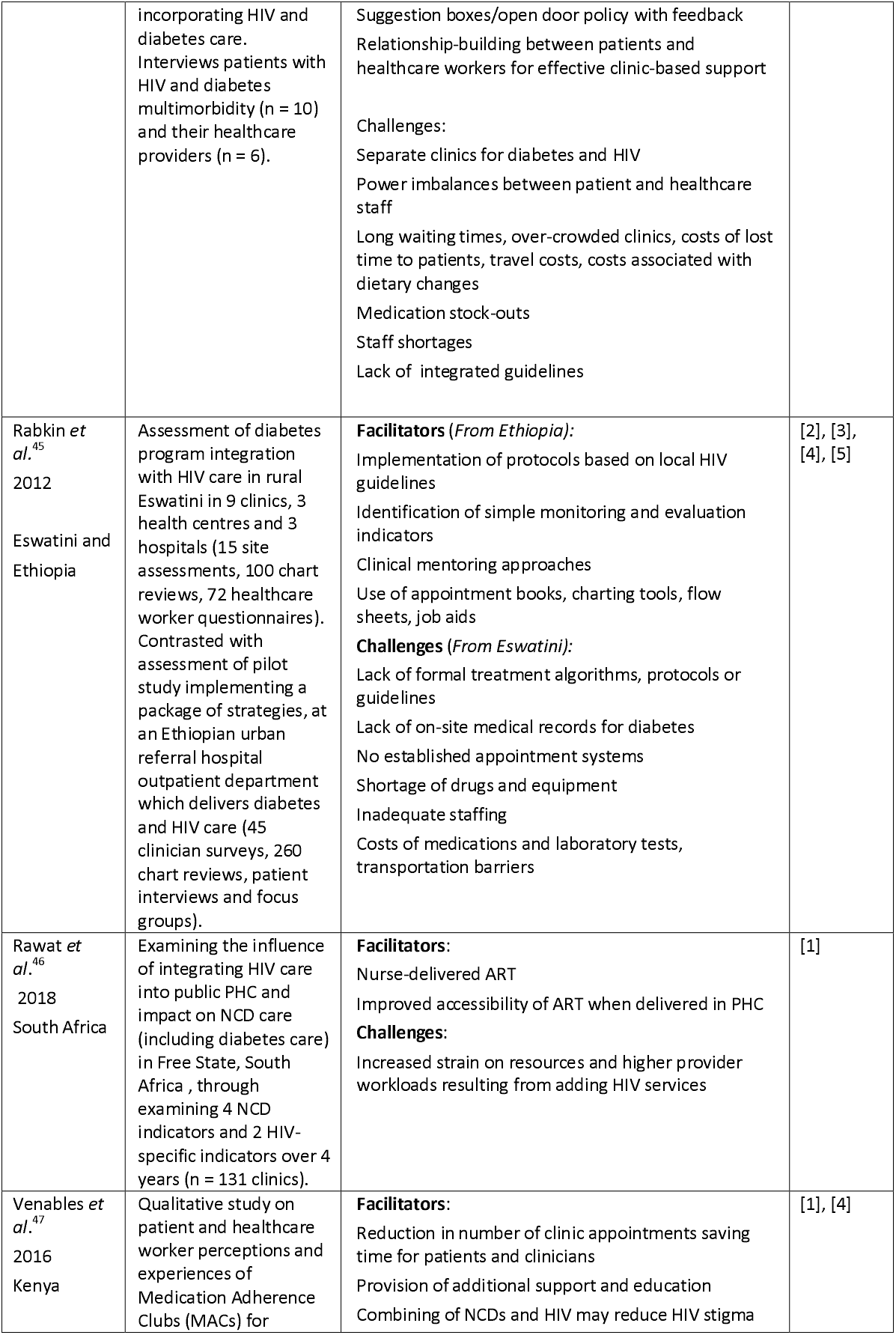
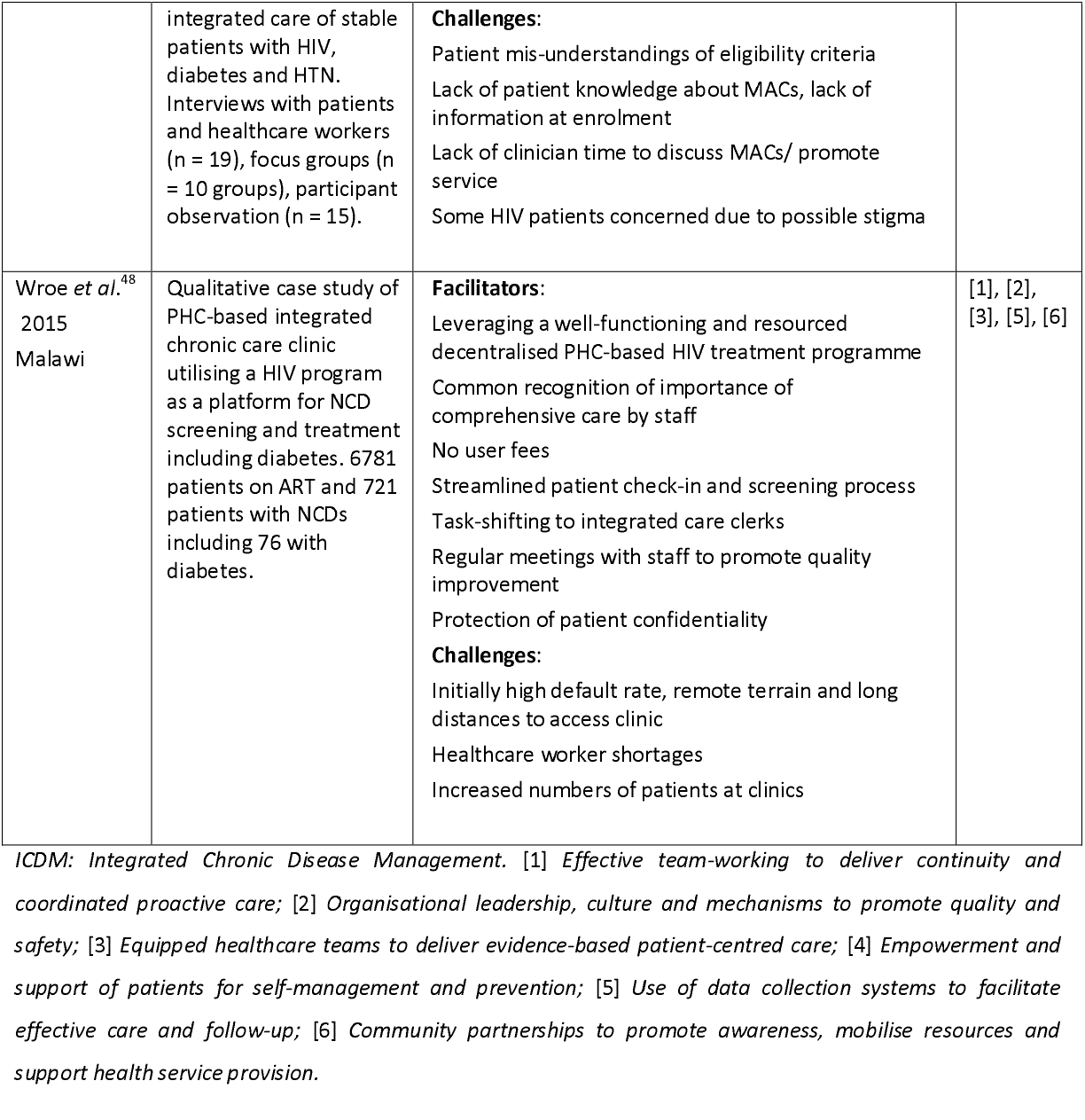
Summary of studies included in a priori framework analysis and identification of a priori themes

| Author, year, country | Description of project/intervention | Highlighted facilitators and challenges to integration | *A priori* themes |
| --- | --- | --- | --- |
| Ameh *et al.*<sup>37</sup><br>2017<br>South Africa | Qualitative case study assessing ICDM framework implementation in 7 facilities incorporating HIV, diabetes and hypertension care. 8 focus groups with patients (n = 56). Interviews with managers (n = 8). | **Facilitators:**<br>Stigma reduction through integration of chronic disease care.<br>**Challenges:**<br>Staff shortages,<br>malfunctioning equipment,<br>process of prepacking medication,<br>long waiting times,<br>transportation costs,<br>drug stock-outs,<br>rigid clinic appointment system,<br>sub-optimal defaulter tracing,<br>stigmatisation from home visits by carers,<br>unprofessional behaviour of nursing staff. | [1], [2],<br>[3], [4] |
| Angwenyi *et al.*<sup>38</sup> | Qualitative analysis examining the implementation of | **Facilitators:**<br>Integrated information system, | [3], [4],<br>[5], [6] |
| 2020<br>Malawi | Malawi's National Health Sector Strategic Plan II, NCD action plan and HIV clinical guidelines for integration of HIV and NCDs including diabetes (DM), at a district PHC level in government and NGO-sponsored facilities. Document search and analysis, structured interviews with managers and providers (n = 15) and NGOs (n = 5). | Volunteer community-based structures.<br>**Challenges:**<br>Disparities in service access<br>Diabetes drug stock outs<br>Lack of clinical space<br>Health workforce shortages<br>Lack of national guidelines for NCD management. |  |
| Chamie *et al.*<sup>39</sup><br>2012<br>Uganda | Analysis of Community Health Campaign for integrated HIV and NCD rapid community screening and linkage to care at 3 months for patients identified to have HIV or diabetes either at local PHC clinic for HIV or at a district hospital clinic for diabetes treatment (n = 2323). | **Facilitators:**<br>Employment of local community members for campaign<br>Efficiency gains of screening for multiple diseases in same campaign<br>HIV treatment services available at local PHC clinic<br>HIV testing is normalised as a routine part of health care, thus reducing stigma<br>Point-of-care (POC) testing<br>**Challenges:**<br>Limited capacity for diabetes treatment at local clinic, as only available at district hospital.<br>Linkage to care after referral was low (61% of newly diagnosed diabetes cases and 34% for HIV).<br>Stigma and transport costs reported as reasons for not linking to care | [3], [6] |
| Davis *et al.*<sup>40</sup><br>2013<br>Botswana | Qualitative case study of elements of general primary care package delivered to HIV positive adults at a PHC clinic incorporating screening and management for NCDs including diabetes, and known local resources and challenges to implementation. | **Facilitators:**<br>Use of nationally available evidenced-based guidelines for management of HIV and diabetes<br>Checklist and ongoing education to support provider adherence to guidelines<br>HbA1c monitoring<br>Diabetic education and counselling by a diabetes specialist nurse available<br>Secondary care referral pathway<br>Diabetes medications available<br>**Challenges:**<br>Glucometers out of stock | [1], [3], [4] |
| Edwards *et al.*<sup>41</sup><br>2015<br>Kenya | Retrospective cohort study evaluating outcomes among people living with HIV (PLHIV) and HIV negative patients attending an NGO- | **Facilitators:**<br>Use of clinical officers and nurses as main caregivers<br>Standardised monitoring and treatment protocols<br>Earlier detection of diabetes in those with HIV<br>**Challenges:**<br>Higher proportion of women in cohort (due to focus | [1], [3] |
|  | operated PHC clinic in an informal settlement in Nairobi providing integrated HIV and chronic disease care including diabetes care (n = 2206). | of NGO services)<br>Lack of health care providers trained in NCD management<br>Lack of relevant national guidelines,<br>Cost of medications |  |
| Frieden *et al.*<sup>42</sup><br>2020<br>Zimbabwe | Qualitative case study evaluation of a nurse-led approach to the integration of diabetes and hypertension (HTN) care with HIV care in an integrated chronic care clinic, or with general outpatient care (including HIV care) at a primary care level (n = 3094 patients). | **Facilitators:**<br>Built on well-functioning HIV infrastructure<br>Context adaptation of guidelines and ongoing mentoring supported by NGO specialists,<br>No user fees<br>On and off-site decision support<br>Regular review meetings<br>Merging of patient records for HIV and NCDs enabled synchronisation of appointments and monitoring and evaluation<br>POC machines for HbA1c measurement<br>Referral system for complex cases<br>Patient empowerment through individual and group education and counselling<br>**Challenges:**<br>Healthcare workforce shortages<br>High patient demand at some sites due to lack of fees<br>Long travel times, staff shortages and high patient volume limited mentorship time<br>Medicines and laboratory reagents supply problems<br>Long laboratory sample transit time, overloaded laboratories<br>Lack of leadership and ownership by staff in some health facilities | [1], [2], [3], [4], [5], [6] |
| Khabala *et al.*<sup>43</sup><br>2015<br>Kenya | Retrospective descriptive study assessing the care of HIV, diabetes and HTN patients enrolled into integrated Medication Adherence Clubs (MACs), at informal settlement in Nairobi. MACs met quarterly to confirm clinical stability, have a short health talk and receive pre-packed medications. (n = 1432 patients enrolled in 47 MACs) | **Facilitators:**<br>Flexible timings of MACs<br>Clear protocols in place<br>Clear eligibility criteria for stable patients to be included in MACs<br>Support for stigmatising conditions<br>Reduced clinician workload as high compliance and low loss to follow-up / need for referral back to regular clinic.<br>**Challenges:**<br>Large numbers of stable patients requiring follow-up<br>Prior to MAC introduction- high clinician work load, long waiting times, and high loss to follow-up rate | [1], [5] |
| Matima *et al.*<sup>44</sup><br>2018<br>South Africa | Qualitative study assessing ICDM framework implementation | Facilitators:<br>Educational materials appropriate to age and education levels available at clinic visits<br>Access to advocacy groups | [1], [3], [4] |
|  | incorporating HIV and diabetes care.<br>Interviews patients with HIV and diabetes multimorbidity (n = 10) and their healthcare providers (n = 6). | Suggestion boxes/open door policy with feedback<br>Relationship-building between patients and healthcare workers for effective clinic-based support<br>Challenges:<br>Separate clinics for diabetes and HIV<br>Power imbalances between patient and healthcare staff<br>Long waiting times, over-crowded clinics, costs of lost time to patients, travel costs, costs associated with dietary changes<br>Medication stock-outs<br>Staff shortages<br>Lack of integrated guidelines |  |
| Rabkin *et al.*<sup>45</sup><br>2012<br>Eswatini and Ethiopia | Assessment of diabetes program integration with HIV care in rural Eswatini in 9 clinics, 3 health centres and 3 hospitals (15 site assessments, 100 chart reviews, 72 healthcare worker questionnaires). Contrasted with assessment of pilot study implementing a package of strategies, at an Ethiopian urban referral hospital outpatient department which delivers diabetes and HIV care (45 clinician surveys, 260 chart reviews, patient interviews and focus groups). | **Facilitators (From Ethiopia):**<br>Implementation of protocols based on local HIV guidelines<br>Identification of simple monitoring and evaluation indicators<br>Clinical mentoring approaches<br>Use of appointment books, charting tools, flow sheets, job aids<br>**Challenges (From Eswatini):**<br>Lack of formal treatment algorithms, protocols or guidelines<br>Lack of on-site medical records for diabetes<br>No established appointment systems<br>Shortage of drugs and equipment<br>Inadequate staffing<br>Costs of medications and laboratory tests, transportation barriers | [2], [3], [4], [5] |
| Rawat *et al.*<sup>46</sup><br>2018<br>South Africa | Examining the influence of integrating HIV care into public PHC and impact on NCD care (including diabetes care) in Free State, South Africa, through examining 4 NCD indicators and 2 HIV-specific indicators over 4 years (n = 131 clinics). | **Facilitators:**<br>Nurse-delivered ART<br>Improved accessibility of ART when delivered in PHC<br>**Challenges:**<br>Increased strain on resources and higher provider workloads resulting from adding HIV services | [1] |
| Venables *et al.*<sup>47</sup><br>2016<br>Kenya | Qualitative study on patient and healthcare worker perceptions and experiences of Medication Adherence Clubs (MACs) for | **Facilitators:**<br>Reduction in number of clinic appointments saving time for patients and clinicians<br>Provision of additional support and education<br>Combining of NCDs and HIV may reduce HIV stigma | [1], [4] |
|  | integrated care of stable patients with HIV, diabetes and HTN. Interviews with patients and healthcare workers (n = 19), focus groups (n = 10 groups), participant observation (n = 15). | **Challenges:**<br>Patient mis-understandings of eligibility criteria<br>Lack of patient knowledge about MACs, lack of information at enrolment<br>Lack of clinician time to discuss MACs/ promote service<br>Some HIV patients concerned due to possible stigma |  |
| Wroe *et al.*<sup>48</sup><br>2015<br>Malawi | Qualitative case study of PHC-based integrated chronic care clinic utilising a HIV program as a platform for NCD screening and treatment including diabetes. 6781 patients on ART and 721 patients with NCDs including 76 with diabetes. | **Facilitators:**<br>Leveraging a well-functioning and resourced decentralised PHC-based HIV treatment programme<br>Common recognition of importance of comprehensive care by staff<br>No user fees<br>Streamlined patient check-in and screening process<br>Task-shifting to integrated care clerks<br>Regular meetings with staff to promote quality improvement<br>Protection of patient confidentiality<br>**Challenges:**<br>Initially high default rate, remote terrain and long distances to access clinic<br>Healthcare worker shortages<br>Increased numbers of patients at clinics | [1], [2], [3], [5], [6] |

ICDM: Integrated Chronic Disease Management. [1] Effective team-working to deliver continuity and coordinated proactive care; [2] Organisational leadership, culture and mechanisms to promote quality and safety; [3] Equipped healthcare teams to deliver evidence-based patient-centred care; [4] Empowerment and support of patients for self-management and prevention; [5] Use of data collection systems to facilitate effective care and follow-up; [6] Community partnerships to promote awareness, mobilise resources and support health service provision.

### Analysis of primary studies against *a priori* framework themes

The studies were analysed and coded using the *a priori* framework themes. All the *a priori* themes were identified in at least 4 studies (Table 2).

‘Effective team-working to deliver continuity and coordinated proactive care’ was included in 9 studies. Several studies highlighted having team members with defined roles as an important strategy for effective teamworking^41 42 46 48^. Continuity was identified as a particular challenge due to high loss to follow up rates^41 48^, with effective appointment systems found to have an important role in delivering continuity and coordinated care. Patients felt that a rigid and inflexible follow-up appointment system represented a barrier to accessing healthcare^37^, while merging follow-up appointment dates for patients with diabetes and HIV comorbidity was identified to be important for improving continuity outcomes^44 42^.

‘Organisational leadership, culture and mechanisms to promote quality and safety’ was included in 4 studies and was identified as a key factor in the success of projects to integrate HIV and diabetes care^42 48^. Benefits were found from having a motivated leader to drive change, make key decisions, and communicate a vision within the organisation^42 48^. Identifying monitoring and evaluation indicators for quality improvement was also highlighted as important for promoting quality and safety^42 45^.

‘Equipped healthcare teams to deliver evidence-based patient-centred care’ was a dominant theme, found within 9 of the 12 primary studies, and supporting the ICCCF modification of the original CCM ‘decision support’ model element to include equipping of healthcare teams^30^. Common findings from the primary studies were that healthcare facilities lacked basic screening equipment^37 38 40 45^, access to laboratory diagnostic tests^38 45^, and a reliable supply of diabetes drugs^37 38 45^. The development and implementation of local and national evidence-based guidelines and diagnostic and treatment algorithms was found to be important for improving patient care^38 41 42 45^. Several studies highlighted clinical mentorship as a method of providing continuing professional education^38 42 45 48^.

‘Empowerment and support of patients for self-management and prevention’ was a theme identified in 7 of the primary research studies. Self-management was mainly supported through the delivery of health education to patients^37 38 40 45 47^, such as a programme to empower patients with diabetes to take responsibility for their health through education about glycaemic control^42^. From a patient perspective, participants in a South African study emphasised the importance of trusting relationships between patients and healthcare workers to facilitate effective clinic-based support for self-management^44^.

‘Use of data collection systems to facilitate effective care and follow-up’ was identified as a theme present in 5 studies. Integration of HIV and NCD patient data into a single, easily accessible patient file was found to facilitate patient data collection for clinical decision-making and monitoring and evaluation activities to improve team performance^38 42 45 48^. Specific, detailed patient records were identified as enabling nursing staff to improve longitudinal follow-up, quantification of patient demand and accurate medication ordering^42 43^.

‘Community partnerships to promote awareness, mobilise resources and support health service provision’ was a theme identified in 4 studies. In Malawi, community health volunteers were mobilised for HIV and NCD patient support in clinics and in the community^38^, and in Uganda local staff were employed to facilitate a screening campaign^39^. Support from local community leadership was also suggested as a way to increase screening uptake in hard-to-reach populations^39^.

### Identification of new themes from primary study thematic analysis

Thematic analysis of results of primary research studies which could not be mapped to the *a priori* framework identified 5 new themes.

#### Improving patient access to chronic disease care

A new theme that emerged from 7 of the studies was that of improving patient access to chronic disease care. Barriers to patient access identified were long distances to travel to clinic, a lack of public transport, low vehicle ownership^37 39 48^, a loss of income resulting from time away from the workplace attending appointments, very long waiting times, and separate appointments for those with more than one chronic disease^37 44^. Several studies identified the need for decentralisation of chronic care to primary care level to reduce travel time and costs^39 42 48^. Three studies considered interventions to reduce waiting times and increase the flexibility of appointment systems^43 47 48^. To remove the financial barrier of out-of-pocket expenditure, free care and medications were provided at the projects in Malawi and Zimbabwe^42 48^.

#### Task-shifting

Six studies highlighted task-shifting as an important strategy for the delivery of integrated HIV and diabetes care in SSA^38 41 42 43 47 48^. Going beyond implementing defined team roles, task-shifting describes a strategy of transferring tasks to healthcare workers with shorter training and fewer qualifications, or to a healthcare cadre trained specifically to perform a limited task only, with the aim of improving service efficiency and accessibility in resource-constrained settings where physician numbers are low^49 50^. Task shifting was identified as an important factor in delivering decentralised integrated chronic care, with several studies advocating the delegation of administrative tasks and screening to non-clinician healthcare cadres^38 48^ and chronic care management to primary care nurses, nurse aides, and non-clinician health educators^42 43 47^.

#### Clinical mentoring

Clinical mentoring is a strategy that has been endorsed by WHO for supporting the scale-up of HIV care in resource-constrained settings^34^, and has a wide scope beyond providing continuing professional education. In 4 of the primary studies, clinical mentorship was used as a strategy to train, retrain, and support healthcare workers for HIV and diabetes care to facilitate task-shifting, decentralisation of services, and capacity building, as well as providing continuing professional development, assessment and supervision to improve quality of care^38 42 45 48^.

#### Stigma and confidentiality

The theme of stigma and confidentiality was found in 5 of the primary research studies. This new theme goes beyond the ICCCF-derived service delivery strategy of raising awareness of chronic conditions and reducing stigma in the community, to highlight the fundamental importance of considering stigma and confidentiality in the design of primary care services and the ongoing support of patients with stigmatising conditions. Both HIV and diabetes were reported by patients to be stigmatising conditions^44 47^. Fear of stigma arising from these conditions was identified by several studies to be an important concern for patients, affecting their willingness to access care^37 39 48^. Conversely, confidentiality was found to be key in allowing HIV patients to feel comfortable to join an integrated NCD and HIV medication adherence club^47^. Patients and healthcare workers felt that integrated HIV and NCDs treatment reduced the stigma of HIV by treating it as any other chronic condition^47^.

#### Patient-provider partnerships

Patient-provider partnerships is a theme that was identified from 5 of the primary studies and incorporates patient feedback, patient experiences and expectations of care, and patient involvement and consultation in service planning. Patients reported a power imbalance between patients and healthcare workers, feeling they lacked the power to ask questions^44^. Matima *et al*. suggested that the healthcare worker-patient relationship needed to be rebalanced to improve patient satisfaction and quality of care^44^. Similarly, Ameh *et al*. noted a provider-patient disconnect with patients expressing dissatisfaction about the behaviour of clinic staff and a lack of trust in their management, leading to the suggestion that assessment of quality of care should incorporate the user’s experience^37^. Wroe *et al*. involved patients in the initial planning for their project, and Frieden *et al*. in the development of locally adapted chronic disease guidelines^42 48^. Venables *et al*. advised that to ensure services are acceptable to patients, it is important to involve people living with chronic diseases in their design to advise on adaptation to local needs and context^47^.

### Expanded thematic framework

The 5 new themes identified from the primary studies and the 6 *a priori* themes confirmed from analysis of the primary research studies were combined to create a new expanded thematic framework, as detailed in Table 3. The new conceptual model has 7 components: Improve patient access to care; Foster patient-provider partnerships; Ensure patient safety and care quality; Empower patients for self-care; Support delivery of comprehensive evidence-based care; Implement effective care, continuity and coordination; and Develop community partnerships. The 7 model components are further formulated to demonstrate a person-centred approach, with the conceptual model visualised as in Figure 2.

**Table 3.** New chronic care model for integrated primary care of HIV and diabetes in SSA.

| Model component | Person-centred formulation | Healthcare service delivery strategies | Themes from expanded thematic framework |
| --- | --- | --- | --- |
| **Improve patient access to care** | *'I am able to access the chronic disease care I need'* | Routine chronic care delivery is decentralised to a primary care level to improve patient access<br>Streamlined and coordinated processes are developed and implemented to improve efficiency of patient services<br>Available healthcare staff are effectively utilised through rational shifting of tasks and have clearly defined roles and responsibilities for delivering integrated chronic care<br>Strategies to reduce patient out-of-pocket expenditure for care and medications are developed | New theme of 'patient access'<br>New theme of 'task-shifting' |
| **Foster patient-provider partnerships** | *'I partner with my healthcare worker to improve my health'* | Patient-healthcare worker communication that facilitates information exchange and shared decision-making is developed and encouraged<br>Healthcare worker training promotes the importance of patient-centred chronic care<br>Service design and implementation safeguards patient confidentiality and seeks to reduce chronic disease stigma<br>Patients are consulted and involved in the planning and implementation of healthcare services<br>Effective avenues of feedback are provided for patient suggestions, concerns, and complaints<br>Care provided is appropriate to the patient's cultural background | New theme of 'patient-provider partnerships'<br>New theme of 'stigma and confidentiality'<br>*A priori* theme of 'effective team-working to deliver continuity and coordinated proactive care' |
| **Ensure patient safety and care quality** | *'I receive safe, high-quality healthcare'* | Organisational leadership develops a culture that supports and promotes the improvement in quality of patient care<br>Effective strategies are promoted for improving the quality of patient care<br>Information for monitoring and evaluation of team performance is collected and used to drive improvements in patient care<br>Providing quality care for patients is incentivised<br>Quality monitoring and improvement activities are routine among all team members<br>Processes are in place for the review of significant events to improve patient safety | *A priori* theme of 'organisational leadership, culture and mechanisms to promote quality and safety'<br>*A priori* theme of 'use of data collection systems to facilitate effective care and follow-up' |
| **Empower patients for self-care** | *'I am empowered with the information and resources to manage my'* | Patients are empowered to take a central role in managing their care<br>Proactive support is provided for patient self-management and prevention efforts over time<br>Effective self-management strategies are provided to patients | *A priori* theme of 'empowerment and support of patients for self-management and prevention' |
|  | *health'* | Suitable resources are provided to support patient self-management and health prevention |  |
| **Support delivery of comprehensive evidence-based care** | *'My healthcare worker has the resources, knowledge and support to provide me with comprehensive evidence-based chronic disease care'* | Healthcare teams are equipped with necessary supplies, medical equipment, laboratory access, and essential medications<br>Guidelines and diagnostic and treatment algorithms are implemented to support delivery of evidence-based chronic disease care<br>Healthcare workers have access to specialist advice<br>Support, education, training, and retraining is delivered through programmes such as clinical mentoring to facilitate decentralisation of chronic disease care<br>Training for healthcare workers is provided to help promote patient self-management and support behavioural change | *A priori* theme of 'equipped healthcare teams to deliver evidence-based patient-centred care'<br>New theme of 'clinical mentoring' |
| **Implement effective care, continuity, and coordination** | *'My healthcare worker has access to my health information to advise me effectively and to plan and coordinate my ongoing care'* | Useful patient data is collected, organised, stored securely and is accessible to guide patient care<br>Continuity is delivered through planned and proactive follow-up to support evidence-based care for chronic diseases<br>Reminder systems are in place for providers and patients<br>Data is used to identify sub-populations for proactive care<br>Data is used for individual care planning<br>Clinical case management is implemented to meet the needs of complex patients<br>Patient care is coordinated through effective communication and sharing of information between team members as well as between healthcare providers | *A priori* theme of 'effective team-working to deliver continuity and coordinated proactive care'<br>*A priori* theme of 'use of data collection systems to facilitate effective care and follow-up'<br>New theme of 'stigma and confidentiality' |
| **Develop community partnerships** | *'My healthcare provider partners with my community to raise awareness, develop services, and increase support for chronic disease care'* | Community awareness of chronic conditions is raised to reduce stigma<br>Partnerships are formed with community leadership and organisations to support patient access to healthcare<br>Local community resources are identified and mobilised to support screening, prevention, and improved management of chronic conditions<br>Complementary preventative and management services are provided where possible, through mobilising an informal network of providers, such as community health workers and volunteers<br>Patients are encouraged to take part in effective community programmes | *A priori* theme of 'community partnerships to promote awareness, mobilise resources and support health service provision' |

**Figure 2.**
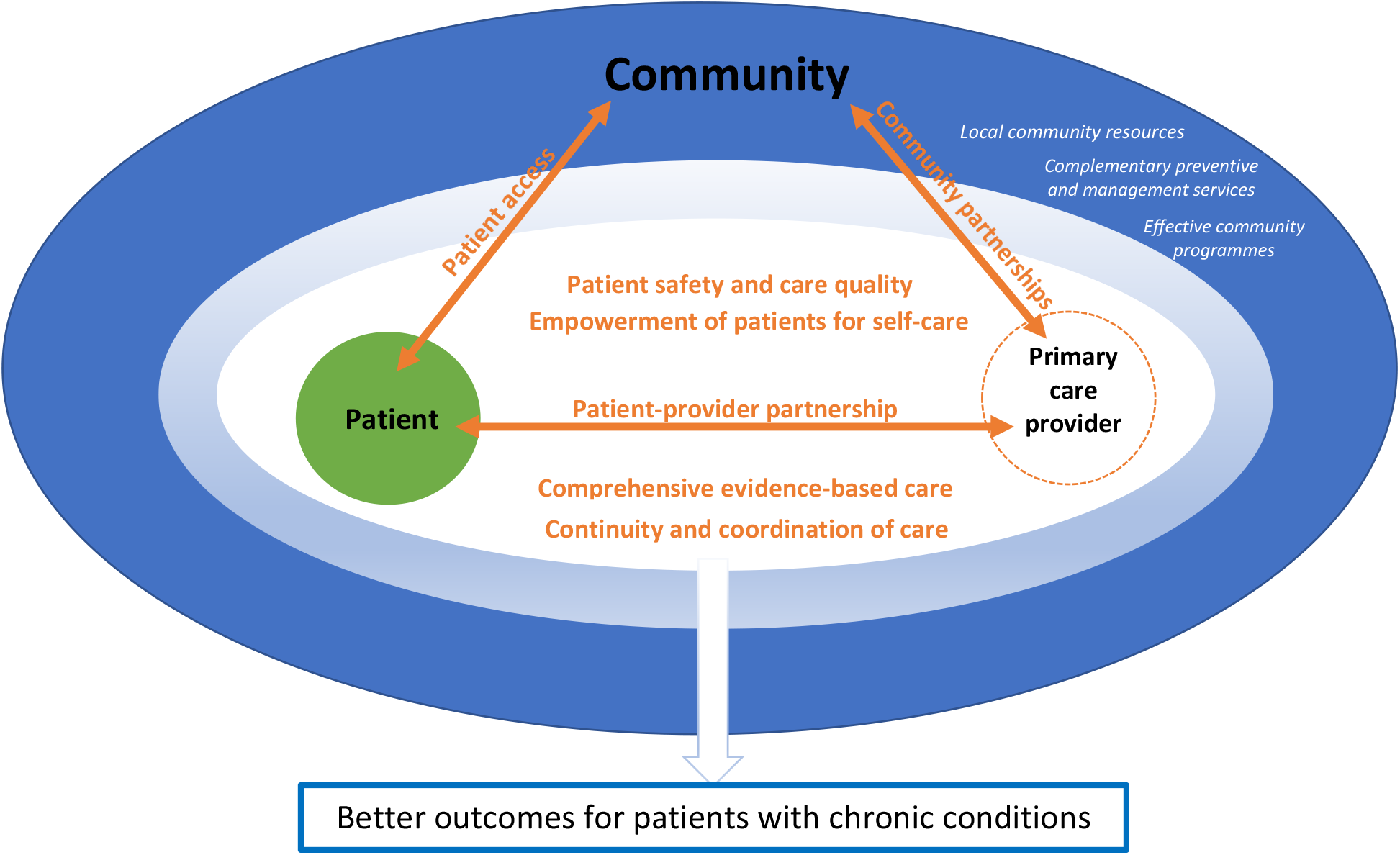
New conceptual model for delivering integrated primary care for chronic diseases incorporating the 7 new model components. Italic text: community resources outside of primary care.

## DISCUSSION

This study has identified chronic care models from which an *a priori* framework for a chronic care model relevant to sub-Saharan Africa (SSA) was produced. Primary research studies investigating integration of HIV and diabetes primary care in SSA were mapped to the *a priori* framework, with new themes identified leading to construction of a novel conceptual model with a person-centred formulation.

The new themes identified from the primary research studies of ‘confidentiality and stigma’, ‘patient-provider partnerships’, and ‘improving patient access’, underline the importance of implementing services which are both acceptable and accessible to patients. These themes have at their heart the real-life struggles and concerns of many patients in SSA and indicate a need for a re-conceptualisation of chronic care models of service delivery from an end-user perspective, demonstrating how the model components relate to improving the care experience of patients living with HIV and diabetes in SSA. The new model incorporates these new themes identified from the primary studies, but also re-orientates the *a priori* themes and service delivery components derived from the CCM and ICCCF to be more person-centred in their formulation (Table 3). Additionally, task-shifting and clinical mentoring are included in the model as key service delivery strategies identified from the primary studies for the contextualisation and practical implementation of a model that delivers person-centred HIV and diabetes care in a resource-constrained SSA setting.

The first aim of the person-centred framing of the new conceptual model components and the person-centred formulation is to provide clarity regarding the aims of the model to policymakers and stakeholders, as well as to the leadership of chronic care programmes and healthcare workers within these programmes. The person-centred framing allows clear linkage of the model components to their outcomes for patients and therefore the reasons for implementing the service delivery strategies to achieve the component goal. If an understanding of the model of care is included within healthcare worker training, this may help with buy-in and motivation for integrated care activities, a problem which was identified in several primary studies as compromising implementation of integrated care^37 42 48^. The second aim of the re-framing is to incorporate the distinctive attributes of primary care, which are access, comprehensiveness, continuity and person-centeredness^51 52^. This links the model more clearly to primary care and underlines its importance as a service delivery approach for provision of integrated, people-centred health services^10^. It has been found that most frameworks used in LMICs for development of primary care services prioritise systems inputs such as human resources, equipment and supplies, facilities, information systems and funding, but fail to give adequate attention to the end goal of person-centred care^52^. The new model aims to address this through re-orientating these important systems inputs around the key functions of service delivery that lead to achieving the outcome of quality, person-centred chronic disease care.

The new model envisages a patient-provider partnership at the heart of chronic disease care, delivering person-centred care that takes account of patient concerns and preferences to create an acceptable service that patients with chronic diseases will want to utilise. Chronic disease care is decentralised, with the primary care provider located within the community of which the patient is a part, to improve patient access. Better outcomes for patients with chronic diseases are achieved by initiatives to increase access to care and deliver the model components through the implementation of the service delivery strategies by the primary care provider. The second partnership is between the primary care provider and the community, to mobilise resources for chronic disease care that patients can access to improve their health outcomes (Figure 2).

The *a priori* themes derived from the CCM and ICCCF models confirm a need to move away from the acute, episodic care common in SSA, towards coordinated and longitudinal care spanning the whole life-course for those with chronic diseases^8 9 10^. Analysis of the primary research studies suggests that integrated care of HIV and diabetes at a primary care level is feasible in a SSA context^53 54^ and can potentially bring benefits such as system synergies from building on established programmes and protocols^42 45^, stigma reduction^47 53^, and increased access to care^48 55 53^.

Each of the *a priori* themes were identified within studies from at least 3 different SSA countries, suggesting that common challenges exist between SSA countries relating to chronic disease care integration, and that the new model for chronic disease care is likely to have relevance across SSA countries. Five of the seven new model components are derived from the original 6 *a priori* framework themes, although they have been re-framed and service delivery strategies reordered to indicate the importance of the component to the end-user through the inclusion of a person-centred formulation (Table 3).

The ‘implement effective care, continuity and coordination’ model component is based upon the *a priori* themes of ‘effective team working’ and ‘use of data collection systems’. The collection of patient data and effective team-working are presented as key strategies to allow continuity in clinical decision-making, care-planning and follow-up, as well as the coordination of care between healthcare workers and providers, which is a pressing need in SSA where fragmented healthcare systems are common^8 56 57 58^. Data security has also been added into this component incorporating an aspect of the new theme of stigma and confidentiality^48^.

The ‘equipped healthcare teams’ *a priori* theme has been re-framed as ‘support comprehensive evidence-based patient care’ to indicate that the aim in adequately equipping health teams with resources, knowledge and specialist support, is to deliver comprehensive and evidence-based HIV and diabetes care^16^. This component also now includes the strategy of clinical mentoring as an approach to facilitate decentralisation, task-shifting and capacity-building in SSA where distance, terrain and lack of healthcare staff pose challenges to equipping staff with knowledge and support^34^.

The model component of ‘patient safety and care quality’ is similar to the *a priori* theme of ‘organisational leadership, culture and mechanisms to promote quality and safety’. This component addresses concerns about the quality of primary care provision in SSA^52 59^ through advocating implementation of robust monitoring and evaluation processes to improve quality and effectiveness of care delivered to patients. Collection of information for monitoring and evaluation of team performance has been included here as an important strategy to drive improvement in the quality of patient care.

Delivery of primary care has been envisaged to involve partnering with people in managing their own health and that of their community^16^. Two new model components address these goals. The first is ‘develop community partnerships’, which corresponds to the *a priori* framework theme of ‘community partnerships to promote awareness, mobilise resources and support health service provision’. Similar to the patient-provider partnership, the community-provider partnership is envisaged as a two-way relationship whereby the community is consulted regarding local health needs and implementation of services, and the community contributes resources, complementary preventative and management services, and human resources to strengthen delivery of chronic disease care^30^. The second model component of ‘patient empowerment for self-care’ is similar to the *a priori* theme of ‘empowerment and support of patients’, and relates to the important role of the healthcare provider in the empowerment of patients for chronic disease self-care^60^. This is facilitated primarily through the patient-provider partnership allowing delivery of patient-centred education and the provision of suitable resources^60 61^, as well as through partnering with community-based informal providers such as health educators and peer-led support groups^30 62 63 64^.

The new themes identified from the primary studies further adapt the new conceptual model for a SSA context. The new model component ‘improve patient access to care’ was derived from the new theme of ‘patient access’. First contact accessibility is a key attribute of primary care^51^, but healthcare workforce shortages, out-of-pocket expenditure, and travel distances to health facilities represent significant barriers to patient access to care in SSA^8 65 66 67 68^. The new model addresses access to care through advocating decentralisation of primary care services and streamlining of processes involved in attending appointments to reduce waiting and travel times and associated indirect costs to patients. Task-shifting is a strategy identified from the primary research studies that has been employed to support improvements in patient access and streamlining of services through the effective utilisation of the limited healthcare workforce in SSA^43 49 69^. Out-of-pocket expenditure also represents a significant barrier to accessing chronic care in SSA^70^. The new conceptual model addresses this through advocating strategies to reduce the direct costs of out-of-pocket expenditures on healthcare^70 71^, ideally, through government financing initiatives, but also through partnering with NGOs and donor organisations where feasible^42 48 54^.

The new model component of ‘patient-provider partnerships’ addresses the issue of acceptability of healthcare provision to patients and focuses on developing person-centred services, incorporating good communication with patients, acknowledgement of patient concerns and preferences, and involvement of patients in decision-making for their care. These elements have the aim of fostering enduring trusting personal relationships with patients to positively impact their care and outcomes^16 44^. Giving patients a voice in decision-making about their care, through feedback about their concerns or complaints, as well as by involvement in identifying local health priorities and the design and implementation of health services, is consistent with WHO’s vision of person-centred primary healthcare^14 16^. The new theme of ‘confidentiality and stigma’ derived from the primary studies, and the service delivery approach of ‘care appropriate to cultural background’ originally from the ‘effective team-working’ *a priori* theme, also feature in this component as service delivery strategies. These strategies aim to improve the acceptability of services, and therefore uptake, through their design and implementation to be culturally sensitive, protect confidentiality and reduce chronic disease stigma^53^.

### Limitations of study

There are several limitations to this study. Firstly, the geographic coverage of the primary studies which cover south and eastern SSA but do not represent any countries in west or central SSA, potentially limiting the generalisability of the new model for the whole of SSA. The BFFS method relies upon availability of primary research studies, and as there have been relatively few studies examining the integration of HIV and diabetes care in SSA^53 54^ it is possible that there are important themes for implementation of integrated chronic disease care in SSA that have not been identified. Also related to the BFFS study design, the service delivery strategies from the *a priori* framework not identified in the primary studies could not confidently be excluded from use in the new conceptual model. This may therefore lead to inclusion of strategies that are not relevant for use in a SSA context. It is also possible that publication bias may lead to failed integration projects not being published, and therefore not included for analysis. The lack of dissonance between the primary studies could be seen as evidence of this, although some studies did record negative findings from implementation projects^20 37 38 42^. Additionally, this study, although outside its stated scope, does not consider policy environment and economic factors that will have a huge influence on the implementation of any model of chronic disease care in SSA^30 31^.

## CONCLUSION

Implementing high-quality chronic disease care in a sustainable way requires moving beyond vertical disease programmes towards integrated healthcare systems, primarily through prioritising investment in primary care. Investing in primary care infrastructure and the healthcare workforce to support decentralisation, and access to person-centred high-quality care for HIV and diabetes, will improve patient outcomes and enable more patients with chronic diseases to lead healthy productive lives. Methods of utilising and training the current healthcare workforce, such as task-shifting and clinical mentoring, can also help to deliver efficient and effective care for patients. This study adds to the available evidence for integrated models of chronic disease care in SSA by building on relevant pre-existing conceptual models, using primary research studies on integration of HIV and diabetes care to develop a new conceptual model relevant to the challenges faced in implementing chronic disease care in SSA. Implementing an integrated model of chronic care that has at its heart the needs and concerns of patients is anticipated to increase utilisation of chronic care services, improve chronic disease care outcomes, and ultimately contribute towards achieving the goal of providing universal high-quality healthcare for all.

## Supporting information

PRISMA 2020 checklist

PRISMA abstract checklist

## Data Availability

All data produced in the present study are available upon reasonable request to the authors

## Contributorship Statement

SH: conception and design of work; acquisition, analysis, and interpretation of data; drafting and revising work; final approval of version to be published. AJ: design of work; interpretation of data; drafting and revising work; final approval of version to be published. Neither author has any competing interests to declare, and no funding was obtained for this work. The review was not registered. There was no patient or public involvement in the design or conduct of this review.

